# Multi-joint approach for assessing lower limb proprioception: reliability and precision in school-aged children

**DOI:** 10.1101/2024.06.19.24308933

**Authors:** Nina Jacobs, Maud van den Bogaart, Ann Hallemans, Pieter Meyns

**Affiliations:** Rehabilitation Research Centre (REVAL), Faculty of Rehabilitation Sciences and Physiotherapy, Hasselt University, 3590 Diepenbeek, Belgium; Research Group MOVANT, Department of Rehabilitation Sciences and Physiotherapy (REVAKI), University of Antwerp, 2610 Wilrijk, Belgium; Research Group for Neurorehabilitation, Department of Rehabilitation Sciences, KU Leuven, 3001 Leuven, Belgium

**Keywords:** Assessment, Proprioception, Joint Position Sense, Reliability, Precision

## Abstract

**Background:** The Joint Position Reproduction (JPR) approach has been commonly used to assess joint position sense (JPS), however, no prior study investigated its psychometric properties in children. This study aimed to assess the reliability and precision of a newly developed multi-joint JPR protocol for assessing lower limb JPS in school-aged typically developing (TD) children.

**Methods:** Ankle, knee and hip JPS was assessed in TD children (aged 5–12 years), on two different days, by a single rater using a standardized JPR protocol (re-identification of a passively placed target position of the ipsilateral joint). The mean and best error(JRE,°) between target and reproduction angle were calculated from three-dimensional(3D) kinematics for each tested joint on both sides for three trials. Furthermore, total, joint- and limb-JRE scores were provided for clinical use. For JPR-reliability, the Intraclass Correlation Coefficient(ICC,2.1) was reported. For JPR-precision, the standard error of measurement (SEM) was calculated.

**Results:** 270 JPR trials were assessed in 15 TD children (8.6±1.2 years,8boys). The mean and best JRE, summarized for all joints for test and retest, was 3.7° and 2.5°, respectively. The ICC were poor to fair(0.01-0.44) for mean JRE, but fair to very good(0.46-0.77) for best JRE. The SEM ranged from 0.8°–1.8°, depending on the joint and side being tested.

**Conclusion:** Evaluating ankle, knee and hip JPS in children, using passive JPR, is more reliable and precise when using the best JRE. This study highlights the need for a multi-joint JPR approach in research and clinics, and provides joint- and limb-specific SEM values.

## 1. Introduction

The development of motor skills, such as walking, running and cycling, requires a integration of the proprioceptive and motor system^[1]^: i.e., to know when and how to generate goal-oriented motor behavior, the central nervous system must have an accurate perception of where the body is located in space and how different body segments are positioned relative to each other. To establish this internal schema, the brain processes input from proprioceptors embedded in the muscles (i.e. muscle spindles), tendons (i.e. Golgi tendon organs), capsuloligamentous structures and skin (i.e., mechanoreceptors)^[2]^. Difficulties in perceiving and integrating this proprioceptive input, resulting from brain damage^[3]^, sport-induced fatigue^[4]^ or injuries^[5]^, particularly in the lower extremities, may diminish a child’s ability to control movements and maintain balance^[6]^. Consequently, this can impact a child’s sport performance, level of competitive success and susceptibility to (re-)injury^[7, 8]^. Also, delayed or deviant proprioceptive development, for example in children with neurodevelopmental disorders^[3, 9]^, can further exacerbate the likelihood of falls and balance problems^[10, 11]^. Moreover, general and sport-specific training programs designed to improve proprioception have shown enhanced injury recovery outcomes and decreased injury rates in various populations, such as young athletes with anterior cruciate ligament injury^[12]^, ankle sprains^[13]^, patellar and Achilles tendinopathy^[14]^ and joint hypermobility syndrome^[15]^. Given its relation to injury and the success of such training programs, the assessment of proprioception has received more attention the past years.

Over the years, researchers have developed a variety of methods to assess different aspects of proprioception (i.e., sense of joint position, movement, trajectory, velocity, force, muscle tension, weight or size)^[16]^. Although other methods are available (e.g., Joint Position Discrimination or Threshold to detection of passive motion^[17]^), the most commonly used, clinically applicable and ecologically valid method to assess proprioception is Joint Position Reproduction (JPR), which focuses on the sense of joint position^[18, 19]^. Herein, proprioception is measured in terms of joint position reproduction error (JRE, in degrees) between the examiner-positioned joint angle and the joint angle repositioned by the subject. The magnitude of JRE is generally accepted as being a useful proprioceptive accuracy indicator (the higher the JRE the lower the accuracy). However, to determine whether meaningful changes in JRE, and thus proprioception, have occurred, but also to ensure reproducibility in repeated trials, knowledge of the measurement error and reliability of JPR method is crucial.

To date, research on the psychometric properties of the JPR method in children is limited^[20]^. Most studies have primarily focused on healthy adults aged 18 or older, demonstrating moderate to good reliability depending on the joint and JPR task being assessed^[21–25]^, However, these reliability findings may not be generalizable to children since proprioception continues to mature until adolescence^[26, 27]^, which causes children to exhibit greater inaccuracy and variability in reproducing joint positions compared to adults^[28]^. This discrepancy could potentially affect the reliability of JPR in children, implicating that reported measurement errors in adults (knee:0.62°^[25]^, hip: 0.63° - 0.72°^[23]^) may not be applicable to children. Furthermore, prior adult’s studies have exclusively focused on single-joint JPR assessments of the more proximal knee^[20–22, 25]^ and hip joint^[23, 24]^, using multi-angle or multi-plane reproduction (i.e. consecutively testing more than one target position or movement plane in the same joint). Generalizability of these reliability findings to other, more distal joints is limited due to the joint-specific nature of JPR testing^[16]^. However, changes in a child’s proprioceptive function resulting from sport-related fatigue, injuries or training, may manifest in one or more joints of the lower limbs, whether or not asymmetric (as previously observed in terms of strength, flexibility, and coordination^[29, 30]^). Performing JPR testing solely on the dominant limb^[20, 21, 24, 25]^, precludes the evaluation of a potential proprioceptive asymmetry between limbs. In summary, a multi-joint JPR approach which includes both proximal and distal joints on both sides of the body, is needed to properly evaluate proprioception in children. However, to the best of the author’s knowledge, no study has reported the reliability and precision of such a JPR approach for assessing lower limb proprioception in children. The aim of the present study was, therefore, to develop a JPR protocol to comprehensively map hip, knee and ankle joint position sense and examine the test-retest reliability and measurement error in typically developing (TD) children aged 5 to 12 years old.

## 2. Methods

A test-retest experimental study design was undertaken to evaluate ankle, knee and hip JPR reliability between two sessions, on two different days with at least four weeks apart to prevent recall bias. This study was conducted between March and August 2022 in accordance to, and approved by the Committee for Medical Ethics (CME) of Antwerp University Hospital (UZA)/University of Antwerp (UAntwerpen), CME of Hasselt University (UHasselt) and the Ethics Committee Research of University Hospital of Leuven (UZ Leuven)/University of Leuven (KU Leuven) (B3002021000145). Five- to twelve-year-old children were recruited from the researcher’s lab environment, an elementary school (Vrije Basisschool Lutselus, Diepenbeek, Belgium), through acquaintances and social media. Written informed consent was obtained from the children’s parents before participation in the study.

## Participants

TD children were included when 1) aged 5 year 0 months until 12 years 11 months, 2) born > 37 weeks of gestation (full term) and 3) cognitively capable of understanding and participating in assessment procedures. They were excluded in case of; 1) intellectual delays (IQ < 70), 2) developmental disorders (e.g. developmental coordination disorder [DCD], Autism Spectrum Disorder [ASD] or Attention Deficit Hyperactivity Disorder [ADHD]), 3) uncorrected visual or vestibular impairments and/or 4) neurological, orthopedic or other medical conditions that might impede the proprioceptive test procedure.

Based on previously reported results on knee JPS in TD children^[20]^, the minimal acceptable level of reliability is 0.39 (H_0:_ ρ_0_=0.39). To provide a desired ICC of 0.80 (H_1_: ρ_1_=0.80), in two-way ANOVA models, with α = 0.05 and β = 0.20, a sample of 15 TD children was needed with n = 2 observations per child, assessed by a single rater^[31]^.

## Data collection and analyses

### Joint Position Reproduction (JPR) protocol

Joint Position Sense was assessed as the child’s ability to re-identify a passively placed target position of the ipsilateral hip (20° of flexion), knee (30° of extension) and ankle joint (15° of dorsiflexion), also known as the passive-ipsilateral JPR protocol (described in Figure 1a).

**Figure 1a.**
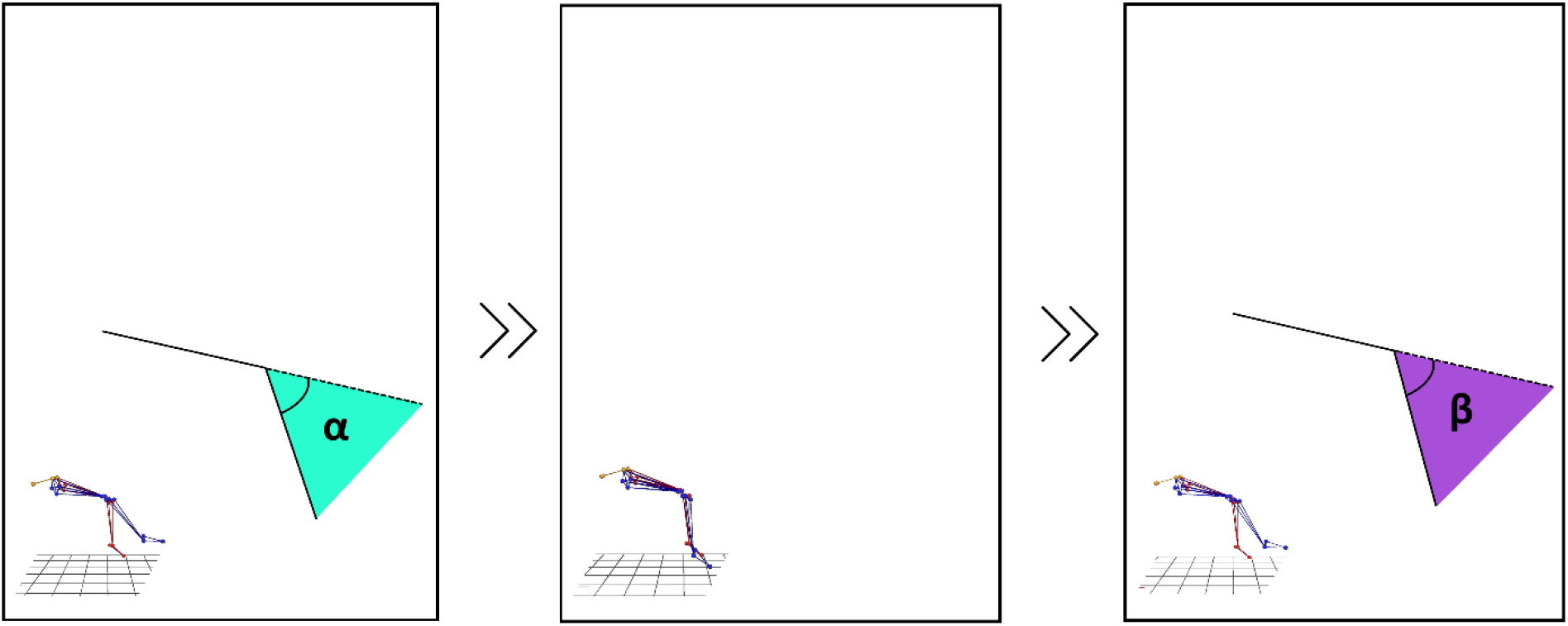
The passive-ipsilateral JPR protocol. **(A) Experience**: from the resting position, the examiner positioned the child’s limb in a predetermined target joint position (hip = 20° of flexion, knee = 30° of extension, ankle = 15° of dorsiflexion) using an inclinometer, distally attached to the moving segment (hip = upper leg, knee = lower leg, ankle = plantar foot side), approximately perpendicular to the flexion-extension axis. **(B) Memory**: after experiencing and memorizing this joint position for 5 seconds, the child’s limb was passively returned to the neutral start position. **(C) Reproduction**: afterwards, the examiner moved the ipsilateral limb back into the same range and the child was asked to re-identify the target joint position as accurate as possible by pressing a button synchronized to motion capture software.

All joints were assessed in isolation on both sides of the body. The dominant limb was defined as the one the child was most comfortable standing on while kicking a ball (representing the limb preferred to perform the balancing aspect of the task). For the ankle and knee JPR task, children were seated blindfolded on the table with the lower legs hanging relaxed and unsupported (90° of knee flexion,), not wearing shoes or socks. The upper leg was fully supported by the table (90° of hip flexion) and hands were crossed on the chest. For hip JPR task, the resting position was the same, only the children sat inclined on the table to align the baseline hip joint angle at 70° of flexion. For the start position of the ankle JPR task, the examiner passively positioned the ankle in maximal plantarflexion. After familiarization (one trial), the ankle, knee and hip JPR tasks were repeated three times for both the dominant and non-dominant leg. The first JPR task undertaken was the knee JPR task, followed by the ankle and hip JPR task respectively. Six trials (3 per body side) were completed randomized (i.e. left or right side) in one joint before moving on to the next joint. In total, within one session, each child had to perform 18 trials (3 trials x 3 tested joints x 2 legs). Performing all JPR tasks took approximately 30 minutes. Depending on the region from which the child was recruited, testing was conducted at the Multidisciplinary Motor Centre Antwerp (M^2^OCEAN, University of Antwerp) or the Gait Real-time Analysis Interactive Lab (GRAIL, Hasselt University).

### Rater

The study was conducted by a registered physical therapist and researcher (NJ) with JPR protocol expertise, gained through preparatory training sessions and a preliminary pilot study (108 JPR trials). To minimize the impact of rater-related sources of error, the examiner was trained to standardize 1) passive joint positioning using an inclinometer (Dr. Rippstein, Lutry, Switserland), at a constant speed, while minimizing skin contact (e.g. knee JRE task: thumb and index finger touch of the calcaneus) and 2) the verbal instructions given (e.g. knee JRE task: ‘concentrate now on the position of your lower leg in space’). To mitigate observer bias, separate test-retest trial record sheets were used.

### Instrumentation

The absolute joint reproduction error (JRE,°) between the target and reproduction angle (Figure 1b) was calculated from 3D kinematics using laboratory-based optoelectronic motion capture (VICON, Oxford Metrics, Oxford, UK) comprising 10 high-speed (100Hz) infrared cameras. Twenty-six 14-mm reflective markers were placed on the child’s body, according to the International Society of Biomechanics (ISB) lower limb recommendations^[32]^: markers were placed on the lateral and medial malleolus, second metatarsal head, the most lateral point on the border of the lateral tibial condyle, the most medial point on the border of the medial tibial condyle, the tibial tuberosity, the lateral and medial femoral epicondyle, the caput femoris, the anterior superior iliac spine and posterior superior iliac, bilaterally. Hip, knee and ankle joint angles were calculated via Euler angles using Vicon Nexus software (v2.12.1; Vicon Inc). The JRE calculations were performed in MATLAB R2022a.

**Figure 1b.**
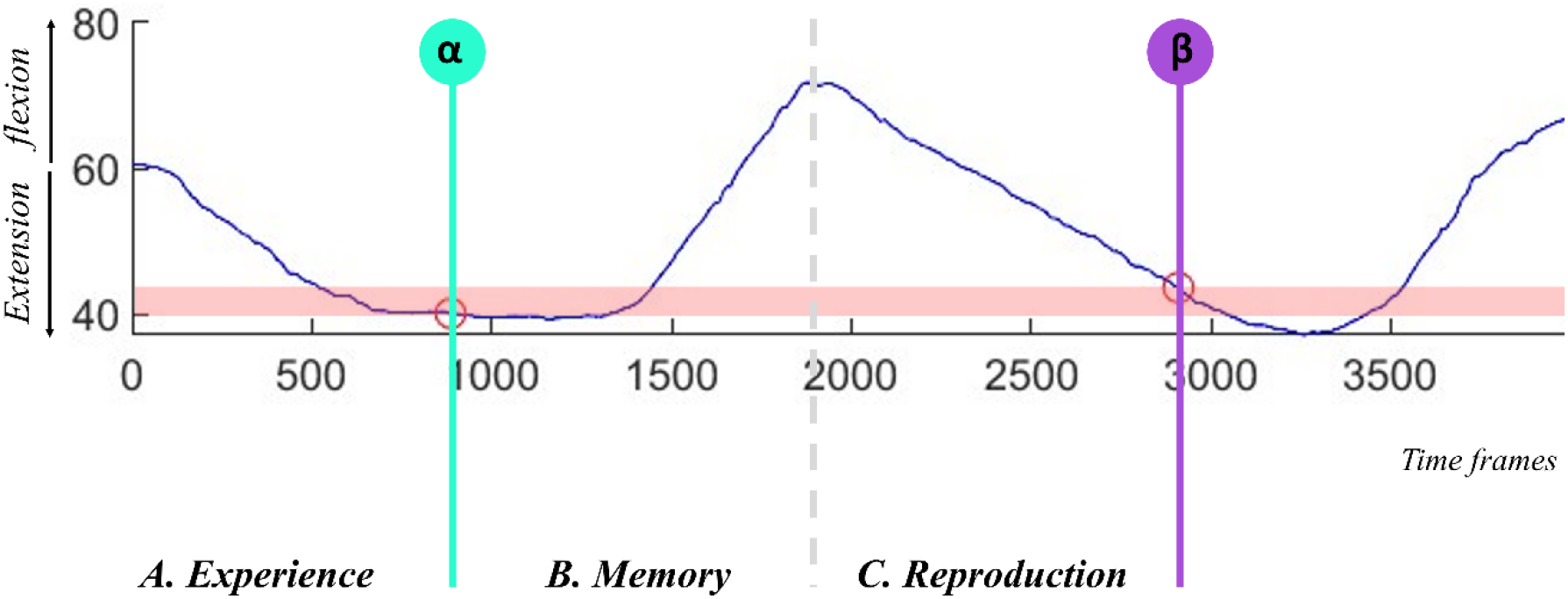
The absolute joint reproduction error (JRE, °) (red) between the passive target (α) and passive reproduction joint angle **(β)** was calculated from 3D kinematics (VICON, Oxford Metrics, Oxford, UK) and used as a measure of JPS. Perfect ankle, knee or hip joint angle reproduction would yield a JRE value of zero.

For each JPR task, JPS was expressed in two different ways:

1. The mean error calculated as the average JRE across the three repetitions, and
2. The best error calculated as the minimal JRE across the three repetitions

Using both expressions of JRE, the following JRE variables were defined:

a. Single-joint JRE of the ankle, knee and hip for the dominant (JREd) and nondominant (JREnd) side separately, resulting in mean and best JREd_ankle_, JREnd_ankle_; JREd_knee_, JREnd_knee_; JREd_hip_, JREnd_hip_;
b. Summed joint JRE of the ankle, knee and hip, summed for the dominant and nondominant side, resulting in mean and best JRE_ankle_, JRE_knee_, and JRE_hip_;
c. Total JRE calculated by adding the summed JRE of the ankle, knee and hip, resulting in a mean and best JRE_total_ (JRE_ankle_ + JRE_knee_ + JRE_hip_)

## Statistical analyses

The normality of the data was evaluated both visually (histogram) and with a 1-sample Kolmogorov-Smirnov goodness-of-fit test.

To define the *test-retest reliability* for the JRE, the Intraclass Correlation Coefficient (ICC) was calculated using a two-way random-effects and absolute agreement ICC model (2,1)^[33]^. Among various types of ICCs, the ICC(2.1) was considered the most appropriate ICC form based on the study design and reliability analyses: 1) “Model” = two-way (subjects x session) - random effects^[34]^, given the JPR protocol is a rater-based clinical assessment where the goal is to generalize reliability results to other trained clinicians; 2) “Definition”= absolute agreement^[35]^, considering both random and systematic sources of errors and 3) “Type” = single rater/measurements, including analyses of single JRE scores of each child for each JPR task, assessed by a single rater. The ICC(2.1) is based upon a two-way repeated measures analyses of variance (ANOVA) with the following formula:

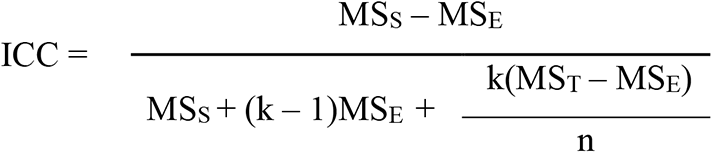

In which MS_S_ is between-subjects mean square, MS_E_ the residual mean square (i.e. random error), MS_T_ the between-session mean square (i.e., systematic error), n the number of children and k the number of observations. This reliability coefficient, or ICC, represents the amount of variance in a set of single JRE scores that is attributable to differences between children (MS_S_) and is interpreted as poor (ICC < 0.40); fair (ICC = 0.40–0.59); good (ICC = 0.60–0.74) or excellent (ICC ≥0.75)^[36]^. To further define the sources of variance for ICC calculations, a two-way repeated measures ANOVA (Sessions [test and retest] x Subjects) was performed for each JRE variable with the significance level (α) set to 0.05.

*The precision* of the JRE was estimated through the standard error of measurement (SEM) defined as:

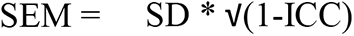

In which SD is the pooled standard deviation (weighted average of standard deviations) for the two measurement sessions.

The ICC and SEM were defined for the following JRE variables: 1) JREd_ankle_, JREnd_ankle_, JREd_knee_, JREnd_knee_, JREd_hip_ and JREnd_hip_; 2) JRE_ankle_, JRE_knee_ and JRE_hip_, only for the tested joint in which the variability (SD) in JREd and JREnd was not significantly different (paired t-test, P>0.05); and 3) JRE_total_.

To demonstrate the practical application of the SEM, two representative cases were selected from the study sample: one from the youngest age group (5-8 years) and one from the oldest age group (9-12 years). For each JRE variable, the calculated SEM was applied to the individual JRE scores to illustrate the expected range of the true scores, outside which one can be confident that a retest JRE score reflects a real change in performance.

Statistical analyses were performed using IBM SPSS Statistics (29.0.1) for Windows.

## 3. Results

Proprioception was assessed during 270 ankle, knee and hip JPR tests of 15 typically developing children (47% girls) with a mean age of 8.58 ± 1.19 years (range 6.74 – 11.11 years). Thirteen children (87%) were right-leg dominant (Table 1). All data were collected in two University facilities; M²OCEAN laboratory (33%) or GRAIL (67%). The median test-retest time interval was 49 days (interquartile range: 35 – 60 days).

**Table 1.** Population characteristics.

|  | Typically developing children (n=15) |
| --- | --- |
| Age, years<sup>a</sup> | 8.58 (1.19) |
| Height, cm<sup>a</sup> | 136.70 (9.67) |
| Leg length dominant leg, cm<sup>a</sup> | 70.75 (8.06) |
| Leg length nondominant leg, cm<sup>a</sup> | 70.00 (7.62) |
| Weight, kg<sup>b</sup> | 30.33 (4.04) |
| BMI, kg/m<sup>2</sup><sup>b</sup> | 16.29 (2.49) |
| Boys/girls<sup>c</sup> | 8/7 (53%/47%) |

Data are presented as mean (SD)<sup>a</sup>, Median (IQR)<sup>b</sup>, Frequency (%)<sup>c</sup>

Descriptive data of the JRE variables, test-retest reliability (as expressed by ICC) and precision (as expressed by SEM) are presented in Table 2. Two exemplary individual cases with their corresponding observed JRE scores (mean and best error) and the estimated true JRE intervals are reported in Table 3.

**Table 2a.** Descriptive data of the JRE *(mean error)*, test-retest reliability (as expressed by ICC) and precision (as expressed by SEM)

|  |  | N | Day 1 (test) | Day 2 (re-test) | Pooled SD, ° | ICC (95% CI) | Variance components |  |  | SEM |
| --- | --- | --- | --- | --- | --- | --- | --- | --- | --- | --- |
|  |  |  | Mean JRE (SD), ° | Mean JRE (SD), ° |  |  | MS<sub>S</sub>(%) | MS<sub>T</sub>(%) | MS<sub>E</sub>(%) |  |
| TOTAL | JRE<sub>total</sub> | 15 | 25.81 (6.53) | 18.90 (4.53) | 5.62 | 0.40 (-0.11 – 0.77) | 21.88 (40) | 23.24 (42) | 9.69 (18) | 4.36 |
| Hip FLEX | JRE<sub>hip</sub> | 15 | 7.29 (2.44) | 5.35 (1.46) | 2.01 | 0.02 (-0.33 – 0.40) | 0 (0) | 1.60 (28) | 4.04 (72) | 2.03 |
|  | JREd<sub>hip</sub> | 15 | 3.75 (1.48) | 2.57 (1.02) | 1.27 | 0.16 (-0.49 – 0.29) | 0 (0) | 0.59 (27) | 1.62 (73) | 1.37 |
|  | JREnd<sub>hip</sub> | 15 | 3.54 (1.38) | 2.79 (0.87) | 1.15 | 0.01 (-0.40 – 0.47) | 0.01 (1) | 0.20 (13) | 1.32 (86) | 1.15 |
| Knee EXT | JRE<sub>knee</sub> | 15 | 8.66 (3.21) | 6.89 (2.80) | 3.01 | 0.44 (-0.02 – 0.76) | 4.51 (44) | 1.26 (12) | 4.57 (44) | 2.26 |
|  | JREd<sub>knee</sub> | 15 | 4.33 (2.29) | 3.47 (1.56) | 1.96 | 0.20 (-0.29 – 0.63) | 0.81 (20) | 0.17 (4) | 3.03 (76) | 1.75 |
|  | JREnd<sub>knee</sub> | 15 | 4.33 (1.95) | 3.42 (1.89) | 1.92 | 0.32 (-0.16 – 0.69) | 1.26 (32) | 0.25 (6) | 2.42 (62) | 1.58 |
| Ankle DF | JRE<sub>ankle</sub> | 15 | 9.86 (3.26) | 6.66 (2.17) | 2.77 | 0.19 (-0.14 – 0.57) | 2.36 (19) | 4.79 (38) | 5.30 (43) | 2.50 |
|  | JREd<sub>ankle</sub> | 15 | 4.93 (2.33) | 3.28 (1.77) | 2.07 | 0.33 (-0.10 – 0.69) | 1.81 (33) | 1.21 (22) | 2.47 (45) | 1.69 |
|  | JREnd<sub>ankle</sub> | 15 | 4.92 (1.73) | 3.38 (1.21) | 1.49 | 0.03 (-0.26 – 0.43) | 0.11 (3) | 1.06 (32) | 2.11 (64) | 1.46 |

Abbreviations: JRE = Joint Reproduction Error; ICC = Intraclass Correlation Coefficient; SEM = Standard Error of Measurement;
Variance components of two-way repeated measures ANOVA (Sessions [test and retest] x Subjects) correspond to the sources of variance for ICC calculations: Mean Squared error of JRE attributable to differences between subjects, between sessions (systematic error) and within subjects (random error)

**Table 2b.** Descriptive data of the JRE *(best error)*, test-retest reliability (as expressed by ICC) and precision (as expressed by SEM)

|  |  | N | Day 1 (test) | Day 2 (re-test) | Pooled SD, ° | ICC (95% CI) | Variance components |  |  | SEM |
| --- | --- | --- | --- | --- | --- | --- | --- | --- | --- | --- |
|  |  |  | Mean JRE (SD), ° | Mean JRE (SD), ° |  |  | MS<sub>S</sub>(%) | MS<sub>T</sub>(%) | MS<sub>E</sub>(%) |  |
| TOTAL | JRE<sub>total</sub> | 15 | 17.16 (7.12) | 12.79 (4.40) | 5.92 | 0.52 (-0.02 – 0.82) | 22.68 (52) | 8.76 (20) | 12.36 (28) | 1.24 |
| Hip FLEX | JRE<sub>hip</sub> | 15 | 4.24 (1.49) | 3.57 (1.26) | 1.38 | 0.13 (-1.34 – 0.70) | 0.26 (13) | 0.11 (5) | 1.65 (82) | 0.97 |
|  | JREd<sub>hip</sub> | 15 | 2.45 (1.44) | 1.82 (0.84) | 1.18 | 0.36 (-0.10 – 0.71) | 0.55 (36) | 0.14 (9) | 0.84 (55) | 0.95 |
|  | JREnd<sub>hip</sub> | 15 | 1.78 (0.83) | 1.75 (0.91) | 0.87 | 0.11 (-0.46 – 0.59) | 0.08 (11) | 0 (0) | 0.65 (89) | 0.81 |
| Knee EXT | JRE<sub>knee</sub> | 15 | 6.20 (2.88) | 4.80 (2.64) | 2.76 | 0.52 (0.06 – 0.80) | 4.35 (52) | 0.77 (9) | 3.27 (39) | 1.28 |
|  | JREd<sub>knee</sub> | 15 | 3.53 (2.30) | 2.75 (1.36) | 1.89 | 0.32 (-0.18 – 0.70) | 1.17 (32) | 0.15 (4) | 2.39 (64) | 1.56 |
|  | JREnd<sub>knee</sub> | 15 | 2.66 (1.90) | 2.04 (1.59) | 1.75 | 0.46 (-0.02 – 0.77) | 1.45 (46) | 0.09 (3) | 1.63 (51) | 1.29 |
| Ankle DF | JRE<sub>ankle</sub> | 15 | 6.73 (4.68) | 4.42 (2.39) | 3.72 | 0.54 (0.05 – 0.82) | 8.70 (54) | 2.31 (14) | 5.09 (32) | 1.49 |
|  | JREd<sub>ankle</sub> | 15 | 3.35 (2.82) | 2.51 (2.08) | 2.48 | 0.77 (0.42 – 0.91) | 4.97 (77) | 0.28 (4) | 1.19 (18) | 1.18 |
|  | JREnd<sub>ankle</sub> | 15 | 3.37 (2.47) | 1.91 (1.18) | 1.94 | 0.30 (-0.12 – 0.67) | 1.39 (30) | 0.91 (20) | 2.36 (51) | 1.62 |

Abbreviations: JRE = Joint Reproduction Error; ICC = Intraclass Correlation Coefficient; SEM = Standard Error of Measurement;
Variance components of two-way repeated measures ANOVA (Sessions [test and retest] x Subjects) correspond to the sources of variance for ICC calculations: Mean Squared error of JRE attributable to differences between subjects, between sessions (systematic error) and within subjects (random error)

**Table 3a.** Observed and estimated true JRE intervals (*mean error*) for two example cases.

|  |  | Case 1 |  | Case 2 |  |
| --- | --- | --- | --- | --- | --- |
|  |  | JRE, ° | JRE, ° | JRE, ° | JRE, ° |
|  |  | Observed | True (range) | Observed | True (range) |
| TOTAL | JRE<sub>total</sub> | 32.87 | (28.51 – 37.23) | 20.24 | (15.88 – 24.6) |
| Hip FLEX | JRE<sub>hip</sub> | 7.90 | (5.87 – 9.93) | 5.36 | (3.33 – 7.39) |
|  | JREd<sub>hip</sub> | 4.76 | (3.39 – 6.13) | 2.76 | (1.39 – 4.13) |
|  | JREnd<sub>hip</sub> | 3.14 | (1.99 – 4.29) | 2.60 | (1.45 – 3.75) |
| Knee EXT | JRE<sub>knee</sub> | 11.44 | (9.18 – 13.70) | 7.32 | (5.06 – 9.58) |
|  | JREd<sub>knee</sub> | 5.53 | (3.78 – 7.28) | 3.20 | (1.45 – 4.95) |
|  | JREnd<sub>knee</sub> | 5.91 | (4.33 – 7.49) | 4.12 | (2.54 – 5.70) |
| Ankle DF | JRE<sub>ankle</sub> | 13.53 | (11.03 – 16.03) | 7.56 | (5.06 – 10.06) |
|  | JREd<sub>ankle</sub> | 7.46 | (5.77 – 9.15) | 3.75 | (2.06 – 5.44) |
|  | JREnd<sub>ankle</sub> | 6.07 | (4.61 – 7.53) | 3.81 | (2.35 – 5.27) |

Abbreviations: JRE = Joint Reproduction Error (in degrees); Case 1 = young child (5-8 years), Case 2 = older child (9-12 years) -

**Table 3b.** Observed and estimated true JRE intervals (*best error*) for two example cases.

|  |  | Case 1 |  | Case 2 |  |
| --- | --- | --- | --- | --- | --- |
|  |  | JRE, ° | JRE, ° | JRE, ° | JRE, ° |
|  |  | Observed | True (range) | Observed | True (range) |
| TOTAL | JRE<sub>total</sub> | 17.85 | (16.61 – 19.09) | 11.05 | (9.81 – 12.29) |
| Hip FLEX | JRE<sub>hip</sub> | 4.18 | (3.21 – 5.15) | 2.95 | (1.98 – 3.92) |
|  | JREd<sub>hip</sub> | 2.27 | (1.32 – 3.22) | 1.18 | (0.23 – 2.13) |
|  | JREnd<sub>hip</sub> | 1.91 | (1.10 – 2.72) | 1.77 | (0.96 – 2.58) |
| Knee EXT | JRE<sub>knee</sub> | 5.85 | (4.57 – 7.13) | 3.80 | (2.52 – 5.08) |
|  | JREd<sub>knee</sub> | 3.08 | (1.52 – 4.64) | 1.75 | (0.19 – 3.31) |
|  | JREnd<sub>knee</sub> | 2.77 | (1.48 – 4.06) | 2.05 | (0.76 – 3.34) |
| Ankle DF | JRE<sub>ankle</sub> | 7.82 | (6.33 – 9.31) | 4.30 | (2.81 – 5.79) |
|  | JREd<sub>ankle</sub> | 4.40 | (3.22 – 5.58) | 1.93 | (0.75 – 3.11) |
|  | JREnd<sub>ankle</sub> | 3.42 | (1.80 – 5.04) | 2.37 | (0.75 – 3.99) |

Abbreviations: JRE = Joint Reproduction Error (in degrees); Case 1 = young child (5-8 years), , Case 2 = older child (9-12 years)

### Mean error

The JRE_total_ was 25.81° (SD 6.53°) and 18.90° (SD 4.53°) for test and retest session, respectively (Table 2a). For all tested joints, regardless of the tested side, the JRE improved (decreased) at retest assessment by 0.75° to 1.65°

The ICC ranged from 0.01 to 0.44 for all JRE variables, indicating poor to moderate test-retest reliability. This implies that an estimated 56% to 99% of observed JRE variance is attributable to within-subjects variability, with the majority of variance components being random (MS_E,_ 43%-86%) rather than systematic (MS_T,_ 4%-38%) (Table 2a).

Overall, the SEM value reported for JRE_total_ was 4.36° indicating that a child’s true JRE summarized for all tested joints being ± 8.55° at 95% confidence (± 1.96 x SEM). As the within-session variability between JREd and JREnd was not significantly different for all joints (ankle: p=0.34, knee: p=0.69; hip: p=0.28), the SEM was also calculated separately for JRE joint scores (JRE_ankle_= 2.50°, JRE_knee_= 2.26° and JRE_hip_= 2.03°) (Table 2a). The SEM for JREd and JREnd of the ankle, knee, and hip varied from 1.15° to 1.75°, with the highest SEM observed in the dominant leg.

### Best error

The JRE_total_ was 17.16° (SD 7.12°) and 12.79° (SD 4.40°) for test and retest session, respectively (Table 2b). For all joints, the best JRE score on day one improved (decreased) by 1.46° or less at re-assessment on day two, depending on the side and the joint being tested. The ICC for test-retest agreement varied from moderate to very good (0.46 – 0.77) for all JRE variables, except for hip flexion (ICC JRE_hip_= 0.13; JREd_hip_= 0.36; JREnd_hip_= 0.11), knee extension of dominant leg (JREd_knee_= 0.32) and ankle dorsiflexion of nondominant leg (JREnd_ankle_= 0.30). Among the latter JRE variables, 64% to 89% of the observed variance was attributable to within-subject variability with 55% to 89% of variance components considered random (MS_E_) and 0% to 9% considered systematic (MS_T_). For the JRE variables with ICC values ≥ 0.46, ≤ 54% of the observed JRE variance was attributable to within-subject variability (MS_E_: 18% - 51%, MS_T_: 3% - 20%).

The SEM for JRE_total_ was 1.24°. With 95% confidence (± 1.96 x SEM), it can be stated that a child’s true JRE best score, summarized for all tested joints, is ± 2.43°. Separate SEM values were also determined for the summed joint JRE of the ankle (JREankle= 1.49°), knee (JREknee= 1.28°) and hip (JREhip= 0.97°), as there was no significant difference in SD between JREd and JREnd across all joints (ankle: p=0.42, knee: p=0.58; hip: p=0.31). Also, For all other JRE variables, the SEM value was less than 2°, ranging from 0.81° to 1.62°.

### Exemplary individual cases

For the summed joint JRE of the ankle, a younger child showed a mean error of 13.53°, while for the knee and hip, the mean errors were smaller (i.e., 11.44° and 7.90°, respectively). Considering the reported SEM values (JRE_ankle_= 2.50°, JRE_knee_= 2.26°, JRE_hip_ = 2.03°), this child’s estimated true JRE mean score varies from 11.03° to 16.03° for the ankle, 9.18° to 13.70° for the knee and 5.87° to 9.93° for the hip. In an older child, lower JRE mean scores were observed for all three joints (JRE_ankle_= 7.56°, JRE_knee_= 7.32°, JRE_hip_ = 5.36°). In this case, a re-assessment JRE mean score ranging from 5.06° to 10.06° for the ankle, 5.06° to 9.58° for the knee and 3.33° to 7.39° for the hip should be interpreted as random variability (Table 3a). When considering the best error, lower JRE values with smaller 95% confidence intervals were observed in both cases, for all JRE variables. For example, in the younger child, JRE_total_ (17.85°) was determined by only 6.33° to 9.31° JRE for ankle dorsiflexion, 4.57° to 7.13° JRE for knee extension and, to a smaller extent, by 3.21° to 5.15° for hip flexion (Table 3b).

## Discussion

This study evaluated the precision and test-retest reliability of a newly developed multi-joint JPR protocol for assessing lower limb JPS in typically developing children aged 5 to 12 years. Our findings indicated that evaluating ankle, knee and hip JPS in TD children is more reliable and precise when using the best score (rather than the mean score). Also, the systematic error between sessions, observed for the best score (0%-20%), is half of the mean score (4%-42%). This implies more consistency in JPR testing across sessions, and thus less tendency to overestimate or underestimate the true JRE score when using the best score. Additionally, total and joint- and limb-specific SEM values were provided for clinical use. These values can be used as an estimated error of a single JPR measurement, accounting for the inherent variability in the JPR measurement process in TD children. This provides clinicians with 95% confidence that a child’s true JRE score lies between two SEMs from the observation made (as described in case reports). For instance, changes in ankle, knee and hip JRE of less than 1.75° should be interpreted with caution before concluding that, due to training or injury, true improvement or deterioration in a child’s ankle, knee or hip JPS has occurred. Understanding and reporting SEM values are therefore crucial when interpreting individual scores or making (clinical) decisions based on JPR assessment.

Given the importance of proprioception in developing motor skills^[1]^, improving motor learning^[37]^ and achieving athlete success^[7, 8]^, proprioceptive assessment received more attention the past years. Although JPR is feasible and commonly used in children^[38]^, there is limited research on its psychometric properties, especially for non-robotic assessments. Only one prior study reported the reliability of JPR for evaluating knee JPS in TD children (ICC: 0.26 – 0.39)^[20]^. Our findings, characterized by ICC values ranging from 0.20 to 0.32, align with those reported by Fatoye et al. (2008). This highlights the challenge of achieving reliable knee JPR assessments, particularly when utilizing mean JRE scores. Children’s attention spans, known to be limited^[39]^ and developing until the age of 10^[40]^, might be challenged in repeated trials. Although it is recommended to implement multiple trials per target position^[41, 42]^, one might counteract for this potential concentration effect by selecting the best JRE instead of calculating the average. Our study supports this hypothesis by showing enhanced reliability for JPR testing with the JRE expressed as the best score (ICC: 0.32-0.46). Besides, Fatoye et al. (2008) focused exclusively on knee JPS assessment. Generalizability to other lower limb joints is thereby limited, as JPR testing is characterized by both joint- and limb-specificity^[16]^. Besides knee proprioception, ankle and hip proprioception can also be altered by sport-related injuries^[5]^, sport-induced fatigue^[4]^ and general or sport-specific training^[43]^, all of which may subsequently lead to altered postural control. This highlights the importance of a comprehensive assessment, measuring (potential deficits in) multiple joints involved in the proprioceptive ability underlying postural control and sport performance. Additionally, assessing both sides is also of importance, as this allows for the evaluation of inter-limb asymmetry (e.g. proprioceptive performance of skill limb [JREd]/the stance limb [JREnd] x 100%)^[44]^, which should also be taken into consideration when assessing performance and injury risk in children engaged in sports^[45]^. By providing a multi-joint JPR approach with total- and joint-specific SEM values, this study allows researchers and clinicians to evaluate changes in a child’s general proprioceptive function (e.g. JRE_total_ > SEM_total_), resulting from improvement or deterioration in one or more joints (e.g. JRE_ankle_ > SEM_ankle,_ JRE_knee_ > SEM_knee,_ JRE_hip_ ≤ SEM_hip_), whether or not asymmetric (e.g. JREd_ankle_ > SEMd_ankle,_ JREnd_ankle_ ≤ SEMnd_ankle_).

However, the low reliability of knee JPR tests (ICC: 0.20 – 0.32) obtained in our study, using the mean JRE scores, are in contrast with previously reported results in healthy adults (ICC 0.42-0.80)^[22, 23, 25]^. This discrepancy could be explained by variations in i) target positions, with adult studies involving more end range positions (40°-70° extension) compared to the midrange position (30° extension) in our study, and ii) positioning – replication procedure, which was actively controlled in the adult subjects, whereas the JPR protocol in our study was passive. Since joint mechanoreceptors and muscle spindles are more sensitive during end range^[46]^ and active movements^[47]^, respectively, it is plausible that these proprioceptors were more activated, which could enhance the accuracy and reliability observed in adults. However, unlike Suner-Keklik et al. (2017) (2.3°)^[25]^, other adult studies did not report better proprioceptive knee accuracy (3.33°-4.65°)^[22, 23]^ compared to our sample of TD children (3.42°-4.33°). Noteworthy is the large between-subject variability in these studies, with ages ranging from 18 to 50 years^[22]^ and from 65 to 81 years^[23]^. This variability could also strongly influence the magnitude of the ICC^[48]^ and may therefore mask poor trial-to-trial consistency in these studies. Moreover, another potential factor contributing to the mixed reliability results could be the participant’s ages. During childhood, periods of rapid growth spurts are often accompanied by musculotendon stiffness^[49]^ and stretching of collagen^[50]^, both of which can affect proprioception^[51, 52]^. Hence, proprioception continues to improve until late adolescence^[53]^, leading to greater variability in reproducing joint positions in children compared to adults^[26]^. This variability may reduce JPR reliability and precision in children, as evidenced by the observed SEM (1.37° - 1.75°), which is larger than those reported in adults for comparable knee (0.63°)^[25]^ and hip JPR procedures (0.63° - 0.72°)^[23]^. This highlights the need for psychometric research on the estimation error of the JPR protocol in children, particularly. Following this, one might also assume that proprioceptive development is marked by an increase in reliability, in addition to the predominantly studied increase in accuracy^[28]^. This may have potentially resulted in an over- and underestimation in SEM for the younger (case 1) and older children (case 2), respectively. To better understand these changes in JRE accuracy, reliability and precision with age, further longitudinal research on ankle, knee and hip JPR in a larger TD sample is needed.

Among different set-ups for the JPR protocol^[18, 22]^, ipsilateral passive reproduction of midrange target positions in sitting was considered the most preferable protocol in this study. Passive JPR, particularly at midrange, enhances its clinical applicability in children, where disease- or injury-related pain, muscle weakness, motor control and/or range of motion limitations can impede active (end range) joint reproductions. Although some studies suggest greater external (ecological) validity^[16]^ and reliability^[22]^ for active JPR end range, smaller passive movements elicit similar muscle spindle stretch responses^[54]^ and midrange positions also achieve reliable results for JPR assessment^[25]^. On the other hand, a seated position precludes the balance challenges and co-contraction of the lower limbs associated with weight-bearing^[55]^, which could also influence a child’s outcome on JPR test.

In this study, potential sources of JRE variance are categorized into two components: systematic bias (e.g., learning or fatigue effects during the test) or random error, due to inherent subject, rater or instrument variation. To mitigate the risk of random test-retest variation, this JPR protocol was standardized as much as possible: same examiner, same verbal instructions and same test conditions with the use of a blindfold and randomized order of target positions (left – right side) for each joint. The relatively small test-retest differences (0.03-1.65°) and between-session variability (MS_T_) observed in JRE, support this test-retest condition control. Additionally, the time between sessions is also comparable to within-subject measurements taken in clinical practice. However, the small improvement in JRE across three joints, within one session, may indicate an overall *learning effect* (JRE_knee_ ≥ JRE_ankle_ > JRE_hip_), as this was the order in which the JPR tasks were carried out. Potential learning effects can be mitigated by also randomizing the order of joints to be tested in addition to randomizing the order of target positions and body side for each joint. Moreover, muscle fatigue has also been reported to adversely affect muscle spindle activity and afferent proprioceptive signalization^[56]^. As a result thereof inaccuracy and variability in reproducing joint positions (and thus JRE error) increases^[57]^. However, the use of a passive JPR protocol precludes a potential *muscle fatigue effect* in this study. Furthermore, contralateral JPR testing^[18]^ and multi-angles reproduction ^[22, 58]^ have been reasoned to require more attention^[59]^, potentially leading to a *concentration fatigue effect*. To reduce this effect and the associated JRE variations, this study opted for a single-angle ipsilateral JPR procedure, particularly as children are known to have a shorter attention span^[39, 40]^. Nevertheless, by assessing multiple repetitions for multiple joints on both sides of the body, the JPR test duration was relatively long (approximately 30 minutes). Consequently, any lapse in concentration may still influence the JRE and pose a risk of greater within-subject variability, which in turns contributes to lower test-retest reliability. To address this potential concentration fatigue effect, our study suggests maintaining multiple trials, as recommended^[41, 42]^, but selecting the best score.

Among all JRE variables, the majority (53% - 96%) of the observed within-subject variance was attributable to random sources of error (MS_E_), such as alertness and attentiveness by the rater. The use of robotic^[60]^ or isokinetic system^[25]^ devices could mitigate rater-related error sources. However, their application in a clinical setting is rather impractical due to their high cost, lack of portability and complexity, particularly in children. Also, the weight of the dynamometer and straps used to stabilize the lower limbs can cause additional sensory input^[61]^. Manual JPR procedures, using the same open kinetic chain positions, are therefore more of interest, as they are also feasible and accurate for children aged 5 years or older ^[60]^. Inter-rater reliability, usually associated with lower ICC and higher measurement error than intra-rater reliability^[48]^, was not assessed. Also, in clinical practice repeated measurements are usually taken by the same trained therapist. Moreover, instrumentation error can also introduce random variations in JRE during repeated testing. To ensure reliability in JPR assessments, it is recommended to measure JPS as the absolute error between the target and reproduction position (JRE), disregarding the direction of the error^[18]^. To quantify these joint positions, laboratory-based 3D optoelectronic motion capture was used. The latter is widely recognized for its reliability and precision in capturing angle, knee and hip kinematics during various tasks, like walking, running and jumping^[62, 63]^. However, variability in marker placement can still be a small but potential source of random error^[64]^. A recently emerging and promising solution to address this issue is markerless motion capture. Using simple video recordings and deep learning, this technique has the potential to analyze kinematics in clinical settings, beyond the laboratory with a high degree of automatization. Despite this, markerless systems may still face challenges in extracting accurate joint centres and angles from recorded video images^[65]^. As such, it remains uncertain whether these techniques are adequately valid (compared to marker-based motion capture), particularly for measuring JRE during ankle, knee and hip JPR tests in children.

## Conclusion

This study was the first to examine the test-retest reliability and precision of a multi-joint approach for assessing lower limb JPS in school-aged TD children. The findings provide valuable insights for clinical assessments of proprioception in children and recommend: i) adopting a comprehensive approach that evaluates both proximal and distal joints for both dominant and nondominant leg, ii) considering the best error as preferred outcome and iii) using the reported total and joint--specific SEM values when making clinical decisions based on JPR assessments. To better understand potential developmental changes in accuracy (JRE), reliability (ICC) and precision (SEM), further longitudinal research on ankle, knee and hip JPR in a larger sample of TD children is warranted.

## Data Availability

All data produced in the present study are available upon reasonable request to the authors

## Acknowledgements

This study was undertaken at Hasselt University and the University of Antwerp, Belgium. Measurements were conducted at the Gait Real-time Analysis Interactive Lab (GRAIL) and the Multidisciplinary Motor Centre Antwerp (M²OCEAN) laboratory, serviced by the university faculties. The authors would like to thank all children and parents who volunteered and participated in this study and the school and master’s students who collaborated and assisted with the recruitment of the children.

